# Elevated meningioma risk among individuals who are Non-Hispanic Black is strongest for grade 2-3 tumors and synergistically modified by male sex

**DOI:** 10.1101/2024.06.13.24308882

**Authors:** Kyle M. Walsh, Mackenzie Price, David R. Raleigh, Evan Calabrese, Carol Kruchko, Jill S. Barnholtz-Sloan, Quinn T. Ostrom

## Abstract

**Background:** Meningioma risk factors include older age, female sex, and African-American race. There are limited data exploring how meningioma risk in African-Americans varies across the lifespan, interacts with sex, and differs by tumor grade.

**Methods:** The Central Brain Tumor Registry of the United States (CBTRUS) is a population-based registry covering the entire U.S. population. Meningioma diagnoses from 2004-2019 were used to calculate incidence rate ratios (IRRs) for non-Hispanic Black individuals (NHB) compared to non-Hispanic white individuals (NHW) across 10-year age intervals, and stratified by sex and by WHO tumor grade.

**Results:** 53,890 NHB individuals and 322,373 NHW individuals with an intracranial meningioma diagnosis were included in analyses. Beginning in young adulthood, the NHB-to-NHW IRR was elevated for both grade 1 and grade 2/3 tumors. The IRR peaked in the seventh decade of life regardless of grade, and was higher for grade 2/3 tumors (IRR=1.57; 95% CI: 1.46-1.69) than grade 1 tumors (IRR=1.27; 95% CI: 1.25-1.30) in this age group. The NHB-to-NHW IRR was elevated in females (IRR=1.17; 95% CI: 1.16-1.18) and further elevated in males (IRR=1.28; 95% CI: 1.26-1.30), revealing synergistic interaction between NHB race/ethnicity and male sex (P_Interaction_=0.001).

**Conclusions:** Relative to NHW individuals, NHB individuals are at elevated risk of meningioma from young adulthood through old age. NHB race/ethnicity conferred higher risk of meningioma among men than women, and higher risk of developing WHO grade 2/3 tumors. Results identify meningioma as a significant source of racial disparities in neuro-oncology and may help to improve preoperative predictions of meningioma grade.

## INTRODUCTION

Meningioma is the most common primary brain tumor in terms of both incidence and prevalence, accounting for >40% of annual primary brain tumor diagnoses in the United States (U.S.).^1,2^ Meningioma incidence increases monotonically from young adulthood through the ninth decade of life,^3^ and ∼1% of the U.S. population will develop a meningioma in their lifetime.^4^ Additional demographic risk factors include higher incidence among women than men, and among African-American individuals than non-Hispanic White individuals.^1,5^ Because meningioma is frequently diagnosed and monitored radiologically, without biopsy or surgical resection, the proportion of tumors classified as World Health Organization (WHO) grade 1, WHO grade 2, or WHO grade 3 varies substantially according to study design.^6^ In registry-based studies, 95% of meningioma is classified as WHO grade 1, with the remaining 5% classified as WHO grade 2 (*i.e*., atypical) or WHO grade 3 (*i.e*., anaplastic).^7^ In tissue-based studies (*i.e*., restricted to patients receiving surgery), up to 20% of meningioma is classified as WHO grades 2-3 using WHO 2021 grading criteria.^8-10^

Epidemiologic studies of meningioma seldom evaluate risk factors in grade-stratified analyses. When such analyses are presented, they are typically split into strata of non-malignant tumors (WHO grades 1-2) and malignant tumors (WHO grade 3). However, recent epigenomic and transcriptomic studies suggest that grade 2 and grade 3 tumors frequently share a similar spectrum of somatic alterations that are not observed in grade 1 tumors.^9,10^ Regardless of grade, meningioma can be a debilitating disease due to morbidity associated with its sensitive intracranial location. Higher-grade meningioma (WHO grades 2-3) frequently display aggressive behavior, with 5-year survival rates as low as 37%.^7^ Further, the neurocognitive sequelae of surgical resection and radiotherapy can be additional sources of morbidity among patients receiving interventional therapy. A recent study observed that 68% of meningioma patients exhibited global neuro-cognitive impairment 18 months after completing their treatment, with 48% unable to return to work.^11^

We recently evaluated the joint effects of biological sex and race/ethnicity on meningioma risk in a large sample of >450,000 U.S. patients.^5^ We observed that the effect of female sex varies significantly across the lifespan, conferring up to 3.5-fold greater risk in women than men during the fifth decade of life. We also observed that, compared to individuals who are non-Hispanic White, individuals who are non-Hispanic Black experience a 1.2-fold higher incidence of grade 1 meningioma and a 1.4-fold higher incidence of grade 2–3 meningioma, whereas Hispanic ethnicity was associated with negligible differences in incidence. Whether the risk of meningioma in African-Americans varies by age and across strata of sex has not been thoroughly evaluated, but could reveal important disparities and inform future strategies for meningioma intervention or tumor interception.

To address this knowledge gap and comprehensively evaluate potential disparities in meningioma incidence among African Americans, we analyzed data from the Central Brain Tumor Registry of the United States (CBTRUS). We examine population-level variation in the incidence of WHO grade 1 and WHO grade 2-3 meningioma among Non-Hispanic Black individuals compared to non-Hispanic White individuals over a sixteen-year period, including the modifying impacts of age and biological sex. Results reveal how age- and sex-specific meningioma incidence rates contribute to racial disparities and further identify high-risk populations that could benefit from targeted approaches for meningioma risk reduction.

## METHODS

### Data Source

Study data are from CBTRUS, which aggregates primary brain tumor incidence data from 52 central cancer registries (47 Centers for Disease Control and Prevention’s [CDC] National Program of Cancer Registries [NPCRs], and 4 National Cancer Institute’s [NCI] Surveillance Epidemiology and End Results [SEERs]).^12^ Collected data capture registered cases from 99.9% of the U.S. population during the time period.^1^ U.S. central cancer registries must collect data on all cancers diagnosed in the United States in accordance with the Cancer Registries Amendment Act (Public Law 102-15), and on all non-malignant primary tumors of the central nervous system (CNS) in accordance with the Benign Brain Tumor Cancer Registries Amendment Act (Public Law 107-260). Study data cover sixteen years, from 2004 (when reporting of non-malignant CNS tumors became mandatory) through 2019, and include all 50 U.S. states and Washington D.C. (excluding data from Nevada for diagnosis years 2018-2019, and tumors diagnosed by autopsy or death certificate only).

Histopathology and behavior codes from International Classification of Diseases for Oncology, 3rd Edition (ICD-O-3) were used to identify meningioma diagnoses and assign WHO tumor grade, as previously described: grade 1 (9530-9534/0, 9537/0), grade 2 (9530-9534/1, 9537-9539/1), and grade 3 (9530-9535/3, 9537-9539/3).^5^ Tumors with ICD-O-3 site codes C70.0 (cerebral meninges) or C70.9 (meninges not otherwise specified) are included in analyses; those with site code C70.1 (spinal meninges) were excluded. Because ∼95% of all meningioma are located intracranially, tumors with site code C70.9 (meninges not otherwise specified) are overwhelmingly intracranial in location and retained in analyses, consistent with prior publications.^5,7^ The WHO grading criteria for meningioma saw revisions in 2004, 2016, and 2021. Our dataset includes only diagnoses prior to 2020, so 2021 WHO revisions cannot affect analyses. Previous sensitivity analyses in 2007-2016 CBTRUS data have revealed minimal impacts of 2004 and 2016 WHO revisions on meningioma analyses, potentially due to rapid uptake of the 2004 revisions and the small number of tumors impacted by 2016 revisions (when brain invasion first qualified a grade 2 designation).^5^ Age, sex, and race/ethnicity data were obtained from the CBTRUS database. Given the study’s aim of expanding prior work on meningioma incidence disparities among African-Americans, analyses are limited to non-Hispanic Black individuals and non-Hispanic White individuals, with the latter having the larger sample size in CBTRUS and serving as a comparator group that maximizes statistical power.

### Statistical Analysis

Population data from the U.S. Census Bureau were obtained from the National Cancer Institute SEER program (http://seer.cancer.gov) and used to calculate incidence rates, age-adjusted to the 2000 U.S. standard population. Average annual age-adjusted incidence rates (AAAIR) with 95% confidence intervals (CIs) were generated using SEER*Stat, by tumor grade (grade 1, grades 2-3), combined race/ethnicity (non-Hispanic White, non-Hispanic Black), biological sex, and 10-year age groups. Incidence rate ratios (IRRs) and 95% CIs were generated using AAAIR, as previously described.^13,14^ IRRs were considered statistically significant at *P* <0.05. Interaction analyses (sex*race/ethnicity) were performed using Poisson models, overall and stratified by age, in R 4.1.3. Figures were created in R 4.1.3.

## RESULTS

Between 2004 and 2019, 56,012 individuals who were non-Hispanic Black and 334,540 individuals who were non-Hispanic White were identified in CBTRUS data as having received a new intracranial meningioma diagnosis. Beginning in the third decade of life (*i.e*., ages 20-29), the AAAIR for meningioma was ∼20% higher among individuals who are non-Hispanic Black than among individuals who are non-Hispanic White (IRR=1.20; 95% CI: 1.12-1.29; P<0.0001) (**Table 1**). This persisted throughout the lifespan, including among those ages 80+ (IRR=1.19; 95% CI: 1.16-1.22; P<0.0001) (**Figure 1**). Interestingly, meningioma incidence was significantly lower among individuals who are non-Hispanic Black as compared to those who are non-Hispanic White in the 0-9 year age group (IRR=0.70; 95% CI: 0.50-0.96; P=0.024) and non-significantly lower in the 10-19 year age group (IRR=0.91; 95% CI: 0.78-1.06; P=0.23). Overall, results indicate that the elevated risk of meningioma in individuals who are non-Hispanic Black begins in young adulthood, peaks in the 60-69 year age group (IRR=1.29), and remains elevated into very old age. Importantly, these incidence patterns were robust to stratification by radiologic versus histopathologic diagnosis (**Supplementary Figure 1**).

**Table 1.** Total cases, average annual age-adjusted incidence rate (AAAIR)^1^, incidence rate ratio (IRR), and 95% confidence interval (CI) for meningioma by 10-year age intervals (CBTRUS: Data provided by CDC’s National Program of Cancer Registries and NCI’s Surveillance, Epidemiology and End Results Program, 2004-2019).

| Age at Diagnosis | Race/Ethnicity | Total<br>(2004-2019) | AAAIR<br>(95% CI) | IRR (95% CI) | P-value |
| --- | --- | --- | --- | --- | --- |
| 0-9 | White non-Hispanic | 247 | 0.07 (0.06-0.08) | REF. |  |
|  | Black non-Hispanic | 48 | 0.05 (0.04-0.07) | 0.70 (0.50-0.96) | **0.0241** |
| 10-19 | White non-Hispanic | 895 | 0.23 (0.21-0.24) | REF. |  |
|  | Black non-Hispanic | 220 | 0.21 (0.18-0.24) | 0.91 (0.78-1.06) | 0.2262 |
| 20-29 | White non-Hispanic | 3,641 | 0.91 (0.88-0.94) | REF. |  |
|  | Black non-Hispanic | 1,072 | 1.10 (1.03-1.16) | 1.20 (1.12-1.29) | **<0.0001** |
| 30-39 | White non-Hispanic | 13,245 | 3.44 (3.38-3.50) | REF. |  |
|  | Black non-Hispanic | 3,287 | 3.89 (3.76-4.02) | 1.13 (1.09-1.18) | **<0.0001** |
| 40-49 | White non-Hispanic | 33,110 | 7.31 (7.23-7.39) | REF. |  |
|  | Black non-Hispanic | 7,579 | 8.65 (8.46-8.85) | 1.18 (1.15-1.21) | **<0.0001** |
| 50-59 | White non-Hispanic | 58,508 | 12.11 (12.02-12.21) | REF. |  |
|  | Black non-Hispanic | 12,230 | 15.47 (15.19-15.751) | 1.28 (1.25-1.30) | **<0.0001** |
| 60-69 | White non-Hispanic | 76,367 | 20.55 (20.40-20.69) | REF. |  |
|  | Black non-Hispanic | 13,073 | 26.52 (26.07-26.98) | 1.29 (1.27-1.32) | **<0.0001** |
| 70-79 | White non-Hispanic | 76,676 | 33.34 (33.11-33.58) | REF. |  |
|  | Black non-Hispanic | 10,811 | 42.34 (41.54-43.15) | 1.27 (1.24-1.30) | **<0.0001** |
| 80+ | White non-Hispanic | 71,851 | 46.96 (46.61-47.30) | REF. |  |
|  | Black non-Hispanic | 7,692 | 55.94 (54.69-57.20) | 1.19 (1.16-1.22) | **<0.0001** |

<sup>1</sup>Rates are per 100,000 population and are age-adjusted to the 2000 US standard population. Abbreviations: CBTRUS, Central Brain Tumor Registry of the United States.

**Figure 1.**
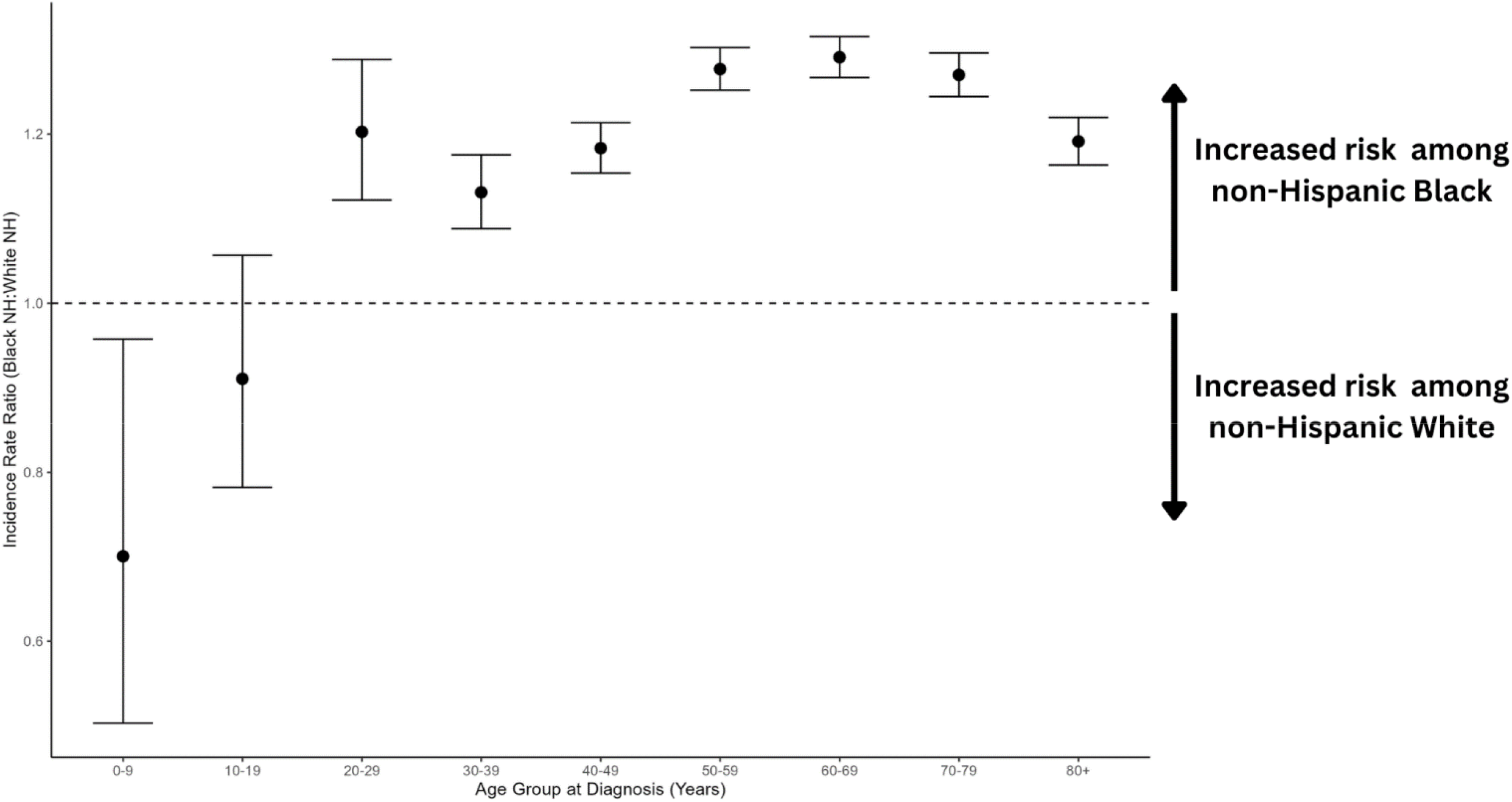
Non-Hispanic Black to non-Hispanic White incidence rate ratios (IRRs) and 95% confidence intervals (CI) for meningioma, by 10-year age group at diagnosis (CBTRUS: Data provided by CDC’s National Program of Cancer Registries and NCI’s Surveillance, Epidemiology and End Results Program, 2004-2019).

Among all 390,552 intracranial meningioma diagnoses in our dataset, 73.0% of cases were diagnosed in females. We next evaluated the IRR in non-Hispanic Black individuals compared to non-Hispanic White individuals, stratified by biological sex, both overall and across 10-year intervals of age at diagnosis (**Supplementary Table 1**). Overall meningioma risk was significantly elevated in non-Hispanic Black females compared to non-Hispanic White females (IRR=1.17; 95% CI: 1.16-1.18; P<0.0001). The magnitude of the meningioma risk associated with non-Hispanic Black race/ethnicity was more pronounced among males (IRR=1.28; 95% CI: 1.26-1.30; P<0.0001), and inclusion of an interaction term (race/ethnicity*sex) revealed the presence of statistically significant multiplicative interaction (P_Interaction_=0.001).

Within 10-year age intervals, we again observed that the elevated risk of meningioma in individuals who are non-Hispanic Black compared to individuals who are non-Hispanic White begins during young adulthood and persists into very old age, but also observed noteworthy differences in these age-specific risks across strata of sex (**Figure 2**). The magnitude of risk associated with non-Hispanic Black race/ethnicity was generally larger among males than among females across age groups, in line with results of the overall interaction analysis. Additional analyses revealed significant multiplicative interaction between sex and race/ethnicity within several 10-year age intervals. Specifically, male sex increased the magnitude of meningioma risk associated with non-Hispanic Black race/ethnicity among each of four 10-year age intervals from 40 years to 79 years of age (**Table 2, Figure 2**). Regression models containing interaction terms require that main effects and interactive effects be interpreted jointly, and provide contrast estimates to aid in model interpretation (**Supplementary Table 2**).

**Table 2.** Meningioma incidence rate ratios (IRR) and 95% confidence intervals (CI) for the joint effects of sex, race/ethnicity, and their interactions from Poisson regression models stratified by 10-year age intervals (CBTRUS: Data provided by CDC’s National Program of Cancer Registries and NCI’s Surveillance, Epidemiology and End Results Program, 2004-2019).

|  | Age 0-9 |  |  | Age 10-19 |  |  | Age 20-29 |  |  | Age 30-39 |  |  | Age 40-49 |  |  | Age 50-59 |  |  | Age 60-69 |  |  | Age 70-79 |  |  | Age 80+ |  |  |
| --- | --- | --- | --- | --- | --- | --- | --- | --- | --- | --- | --- | --- | --- | --- | --- | --- | --- | --- | --- | --- | --- | --- | --- | --- | --- | --- | --- |
|  | IRR | 95% CI | P | IRR | 95% CI | P | IRR | 95% CI | P | IRR | 95% CI | P | IRR | 95% CI | P | IRR | 95% CI | P | IRR | 95% CI | P | IRR | 95% CI | P | IRR | 95% CI | P |
| Age | 0.99 | 0.95-1.03 | 0.7 | 1.15 | 1.13-1.18 | <0.001 | 1.15 | 1.14-1.17 | <0.001 | 1.11 | 1.10-1.12 | <0.001 | 1.06 | 1.06-1.07 | <0.001 | 1.05 | 1.04-1.05 | <0.001 | 1.06 | 1.06-1.07 | <0.001 | 1.04 | 1.04-1.04 | <0.001 | 1.04 | 1.04-1.04 | <0.001 |
| Male Sex | 0.87 | 0.68-1.12 | 0.3 | 0.85 | 0.74-0.97 | 0.013 | 0.55 | 0.51-0.59 | <0.001 | 0.33 | 0.32-0.35 | <0.001 | 0.29 | 0.28-0.29 | <0.001 | 0.34 | 0.33-0.34 | <0.001 | 0.43 | 0.42-0.44 | <0.001 | 0.51 | 0.50-0.52 | <0.001 | 0.62 | 0.61-0.64 | <0.001 |
| Non-Hispanic Black race/ethnicity | 0.49 | 0.29-0.79 | 0.005 | 0.92 | 0.75-1.12 | 0.4 | 1.24 | 1.14-1.35 | <0.001 | 1.09 | 1.05-1.14 | <0.001 | 1.11 | 1.08-1.14 | <0.001 | 1.18 | 1.16-1.21 | <0.001 | 1.22 | 1.19-1.24 | <0.001 | 1.20 | 1.17-1.22 | <0.001 | 1.17 | 1.14-1.20 | <0.001 |
| Male sex * Non-Hispanic Black race/ethnicity | 1.86 | 0.99-3.56 | 0.057 | 0.97 | 0.72-1.31 | 0.8 | 0.88 | 0.76-1.02 | 0.089 | 1.02 | 0.94-1.12 | 0.6 | 1.17 | 1.10-1.24 | <0.001 | 1.22 | 1.17-1.28 | <0.001 | 1.11 | 1.07-1.16 | <0.001 | 1.10 | 1.05-1.15 | <0.001 | 0.97 | 0.92-1.03 | 0.3 |

**Figure 2.**
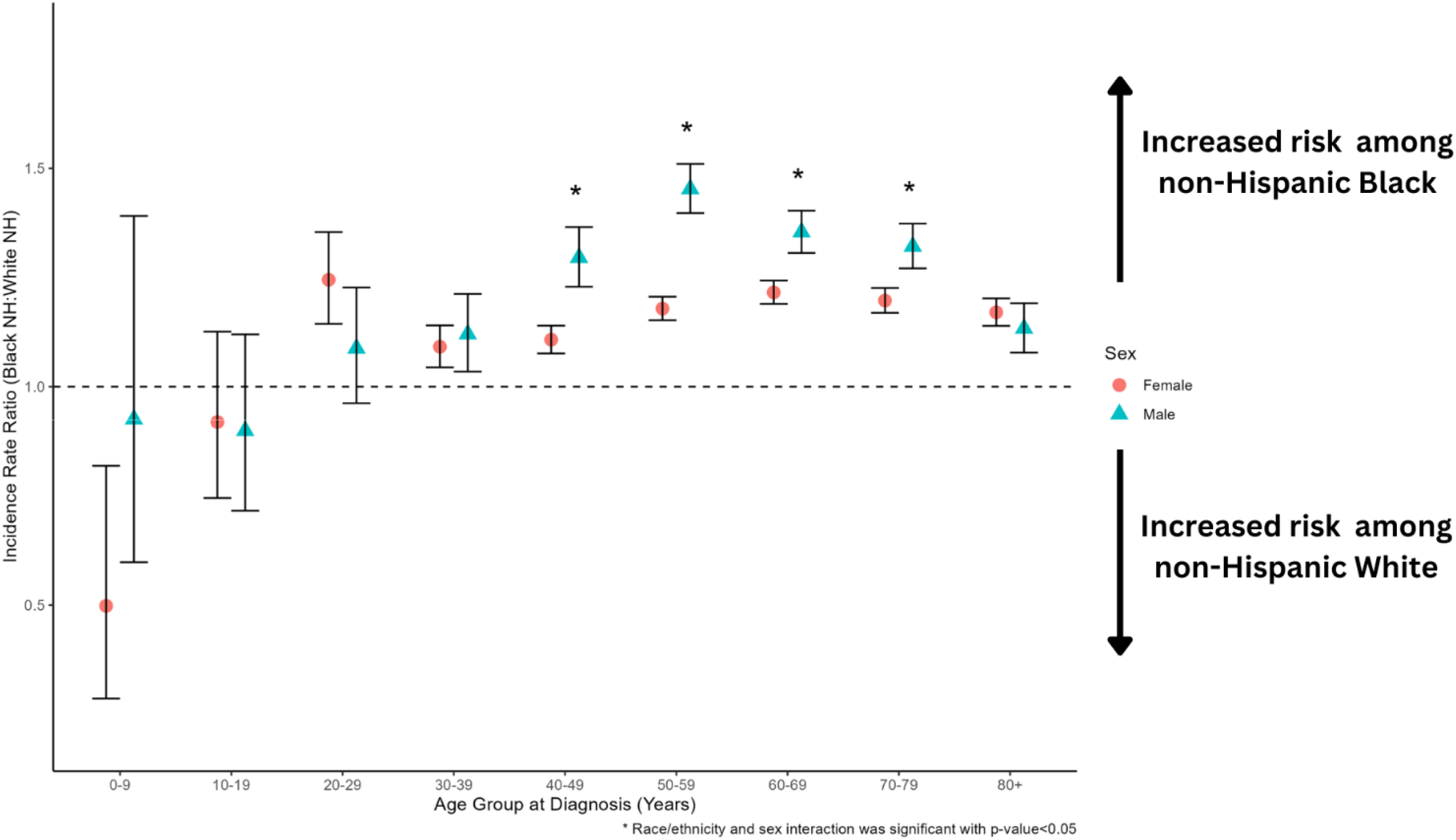
Non-Hispanic Black to non-Hispanic White incidence rate ratios (IRRs) and 95% confidence intervals (CI) for meningioma, by 10-year age group at diagnosis and stratified by biological sex. Asterisks denote statistically significant interaction (P<0.05) in the meningioma risk associated with race/ethnicity among males compared to females (CBTRUS: Data provided by CDC’s National Program of Cancer Registries and NCI’s Surveillance, Epidemiology and End Results Program, 2004-2019).

As implied by interaction analyses, the largest sex-stratified IRR comparing non-Hispanic Black individuals to non-Hispanic White individuals was observed among males, specifically in the 50-59 year age group (IRR=1.45; 95% CI: 1.40-1.51; P<0.0001). The largest IRR comparing non-Hispanic Black females to non-Hispanic White females was more modest and occurred earlier in the lifespan, among the 20-29 year age group (IRR=1.25; 95% CI: 1.14-1.35; P<0.0001). The meningioma risk associated with non-Hispanic Black race/ethnicity among females subsequently declined in the 30-39 year age group (IRR=1.09; 95% CI: 1.04-1.14; P=0.0001), then trended upward in subsequent decades (**Figure 2**).

Among all meningioma diagnoses in our dataset, 5.9% were classified as WHO grade 2 or grade 3. We next evaluated the IRR in non-Hispanic Black individuals compared to non-Hispanic White individuals within 10-year intervals of age and stratified by WHO tumor grade (**Figure 3, Supplementary Table 3**). Beginning in the third decade of life (ages 20-29), risk of grade 1 meningioma was significantly elevated in individuals who are non-Hispanic Black compared to individuals who are non-Hispanic White (IRR=1.25; 95% CI: 1.16-1.33; P<0.0001). The largest non-Hispanic Black to non-Hispanic White IRR for grade 1 meningioma occurred in the 50-59 year age group (IRR=1.27; P<0.0001) and the 60-69 year age group (IRR=1.27; P<0.0001), but was only modestly attenuated in the following decade of life (IRR=1.26; P<0.0001). In analyses of WHO grade 1 tumors, significant interaction between race/ethnicity and sex were again observed during each of four 10-year age intervals from 40 years to 79 years of age (**Supplementary Figure 2**).

**Figure 3.**
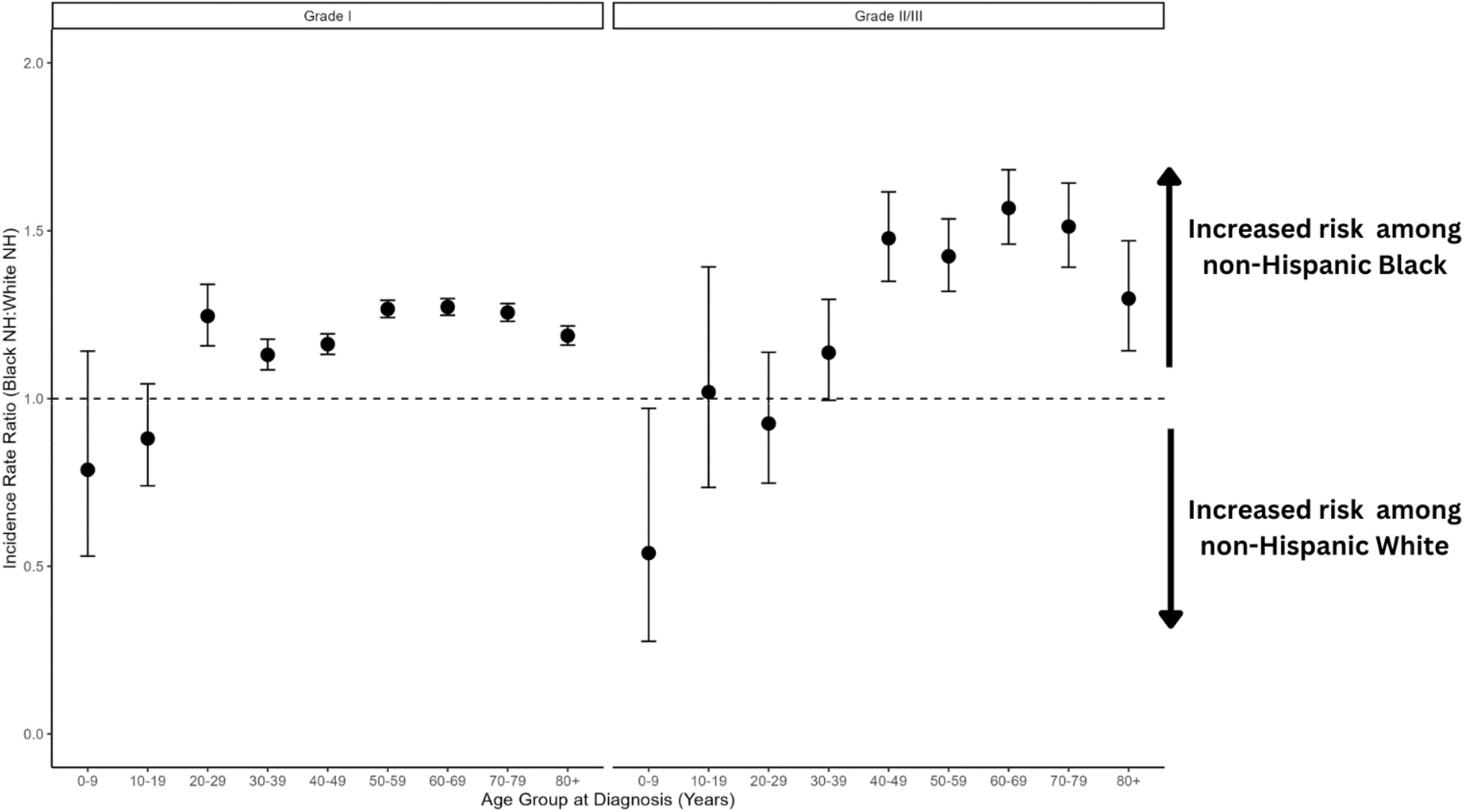
Non-Hispanic Black to non-Hispanic White incidence rate ratios (IRRs) and 95% confidence intervals (CI) for meningioma, by 10-year age group at diagnosis and stratified by central nervous system (CNS) WHO grade (CBTRUS: Data provided by CDC’s National Program of Cancer Registries and NCI’s Surveillance, Epidemiology and End Results Program, 2004-2019).

Non-Hispanic Black race/ethnicity was associated with larger increases in risk of grade 2-3 meningioma than observed for grade 1 meningioma (**Figure 3, Supplementary Table 3**). A non-significant increased risk of grade 2-3 tumors was observed in individuals who are non-Hispanic Black compared to individuals who are non-Hispanic White in the 30-39 year age group (IRR=1.14; 95% CI: 1.00-1.30; P=0.059). The magnitude of effect increased the following decade in the 40-49 year age group (IRR=1.48; 95% CI: 1.35-1.629; P<0.0001) and reached its zenith in the 60-69 year age group (IRR=1.57; 95% CI: 1.46-1.69; P<0.0001). Although interaction analyses had reduced power in age-stratified analyses of WHO grade 2-3 tumors, the IRR associated with non-Hispanic Black race/ethnicity was generally larger in males than in females. Male sex and non-Hispanic Black race/ethnicity displayed significant interaction in their associations with grade 2-3 meningioma risk during three different decades of life, including synergistic interaction in the 10-19 and 50-59 year age groups and antagonistic interaction in the 20-29 year age group (**Supplementary Figure 2**).

## DISCUSSION

We present sixteen years of nationally-representative meningioma incidence data covering 99.9% of the U.S. population. Analysis of 390,552 meningioma diagnoses, including 56,012 individuals who are non-Hispanic Black, reveals how race/ethnicity and biological sex jointly influence meningioma risk across the lifespan. During the first decade of life, individuals who are non-Hispanic Black have a significantly *decreased* risk of meningioma compared to individuals who are non-Hispanic White. However, this relationship is inverted by the third decade of life and meningioma risk remains elevated in African-Americans beyond eighty years of age. We further reveal that the effect of non-Hispanic Black race/ethnicity on meningioma risk is significantly modified by sex. Specifically, the elevated meningioma risk in non-Hispanic Black men relative to non-Hispanic White men is significantly greater than the meningioma risk in non-Hispanic Black women relative to non-Hispanic White women. Additionally, the increased risk of meningioma associated with non-Hispanic Black race/ethnicity varies by WHO tumor grade and was highest for grade 2-3 tumors throughout adulthood.

That female sex and African-American race are associated with elevated meningioma incidence aligns with prior research.^5,7,15^ In partnership with state-level cancer registrars, CBTRUS data comprehensively capture diagnoses in racial/ethnic minority populations and provides unparalleled statistical power for investigating the interactive effects of sex and race/ethnicity within 10-year age intervals. The large sample size, including >22,000 WHO grade 2-3 tumors, also clarifies how the elevated incidence of higher-grade meningioma in non-Hispanic Black individuals relative to non-Hispanic White individuals varies by sex and across the lifespan.

Higher-grade meningioma is a challenging clinical entity with poorly understood epidemiology due to its lower incidence than that of grade 1 meningioma. Prior epidemiologic research on meningioma has grouped WHO grade 1 and WHO grade 2 tumors together for analyses, but this appears inappropriate based on recent epigenomic and transcriptomic analyses revealing minimal molecular overlap between grade 1 and grade 2 tumors, and greater overlap between grade 2 and grade 3 tumors.^6,9,10,16^ The analyses presented here support this analytic approach, revealing that non-Hispanic Black race/ethnicity is associated with an increased risk of grade 2-3 tumors that is also robust to stratification by radiologic versus histopathologic diagnosis.

We previously demonstrated that females experience higher meningioma incidence than males by the second decade of life, with the female-to-male IRR peaking at ∼3.4 in the fifth decade of life, then declining in subsequent decades.^5^ Additionally, up 80% of meningioma tumors express progesterone receptor and 40% express estrogen receptor.^17-19^ These epidemiologic and neuropathologic data support a hypothesized hormonal etiology in meningioma tumorigenesis. Differences in age- and sex-stratified meningioma risk associated with non-Hispanic Black race/ethnicity are intriguing in the context of racial/ethnic variation in hormonal regulation.

A body of prior research reveals that African-American girls experience earlier and more accelerated pubertal maturation than non-Hispanic White girls, as indicated by younger ages at initiation of pubarche, thelarche, and menarche.^20^ However, observed racial/ethnic differences in pubertal timing are in the range of 6-18 months and are therefore unlikely to fully explain elevated meningioma risk in African-American women over 10-year age intervals, as observed in our study. These may, however, partially explain the isolated peak in IRR observed in 20-29 year old females in Figure 2, which was also the lone decade of life during which male sex displayed *antagonistic* interaction with non-Hispanic Black race/ethnicity in analysis of grade 2-3 tumors. Importantly, the time interval between pubarche and menarche also appears to vary by race/ethnicity.^20^ These differences in “pubertal tempo” may have larger impacts on the incidence of hormonally-mediated neoplasms, including meningioma.

Measures of adiposity (*e.g*., body mass index (BMI), free fat mass, waist-hip ratio) affect serum sex-hormone concentrations and are also validated meningioma risk factors.^21-23^ Unfortunately, the registry-based approach used in our analysis is unable to account for individual-level differences in body habitus and will require follow-up in datasets containing either electronic medical records or, potentially, insurance claims. While the effect of race/ethnicity on meningioma risk waned in later decades of life among both males and females, the drop was more precipitous in males. Whether this is biologically meaningful or potentially influenced by a more pronounced healthy survivor bias among elderly men than elderly women will require alternative datasets and analytic approaches that incorporate competing risks.

In addition to ionizing radiation exposure and elevated BMI, other modifiable/exogenous factors conferring meningioma risk include cigarette smoking^24^ and potentially airborne particulate pollution.^25^ Whether such environmental exposures could contribute to elevated meningioma incidence in African-American individuals is unclear, as is the possibility that such factors might operate by altering the hormonal milieu, although menopause is known to occur earlier in smokers.^26^ Epidemiologic studies also indicate that age at menopause varies across racial/ethnic groups, with African-American women experiencing menopause 6-12 months earlier than non-Hispanic White women.^26^ However, other large studies attribute observed racial/ethnic differences in menopause timing to geography, with the effect of race/ethnicity disappearing after adjustment for regional variation within the U.S.^27^ If environmental exposures contribute to the elevated meningioma incidence observed among African-American individuals, it suggests that such risk factors are likely to be distributed differently across U.S. racial/ethnic groups and to be more prevalent where African-Americans constitute a greater proportion of the local/regional population.

Our study has several important limitations. First, meningioma cases in this analysis include radiologic diagnoses and molecular diagnoses. Completeness of data collection for radiologic diagnosis of non-malignant brain tumors varies by state and could contribute to regional variation in meningioma incidence. Further, a modest subset of patients had ICD-O-3 site code C70.9 (meninges not otherwise specified), and thus a small number of individuals included in analysis may have extracranial (*i.e*., spinal) meningioma. Molecular biomarkers have only recently been incorporated into the WHO classification of meningioma (e.g., *CDKN2A* loss) and are not yet available in CBTRUS data.^8^ Finally, the latency period between exposures and meningioma development is likely to differ across strata of tumor grade,^28^ and this may affect how age-stratified incidence rates are interpreted.

While meningioma can be managed without neurosurgical intervention in the majority of patients, a sizeable minority require aggressive therapy.^29^ There is currently no reliable method to determine meningioma grade in the preoperative setting, which represents a major gap in neuro-oncology practice.^6^ While less invasive approaches to determine meningioma grade are emerging,^30-32^ the population-based data presented here indicate that both liquid biopsy and radiomic-based approaches could be improved by incorporating demographic data elements such as age, sex, and race/ethnicity.

Such improvements in discriminative ability have the potential to improve clinical care by minimizing both under-treatment and over-treatment of meningioma. Further research will be needed to translate these findings into cohorts of molecularly-classified meningioma and to explore the contributions of environmental risk factors to racial disparities in their incidence.

## Data Availability

The data used in the present study are provided to the authors under contract with the CDC and are not publicly available

## Acknowledgements

The CBTRUS data were provided through an agreement with the Centers for Disease Control’s National Program of Cancer Registries. In addition, CBTRUS used data from the research data files of the National Cancer Institute’s Surveillance, Epidemiology, and End Results Program, and the National Center for Health Statistics National Vital Statistics System. CBTRUS acknowledges and appreciates these contributions to this report and to cancer surveillance in general. Contents are solely the responsibility of the authors and do not necessarily represent the official views of the CDC or the NCI.

## REFERENCES

1. Ostrom QT, Price M, Neff C, et al. CBTRUS Statistical Report: Primary Brain and Other Central Nervous System Tumors Diagnosed in the United States in 2016-2020. Neuro Oncol. 2023;25(12 Suppl 2):iv1–iv99.

2. Neff C, Price M, Cioffi G, et al. Complete prevalence of primary malignant and nonmalignant brain tumors in comparison to other cancers in the United States. Cancer. 2023;129(16):2514–2521.

3. Walsh KM. Epidemiology of meningiomas. Handb Clin Neurol. 2020;169:3–15.

4. Huntoon K, Toland AMS, Dahiya S. Meningioma: A Review of Clinicopathological and Molecular Aspects. Front Oncol. 2020;10:579599.

5. Walsh KM, Price M, Neff C, et al. The joint impacts of sex and race/ethnicity on incidence of grade 1 versus grades 2–3 meningioma across the lifespan. Neuro-Oncology Advances. 2023.

6. Wang JZ, Landry AP, Raleigh DR, et al. Meningioma: International Consortium on Meningiomas (ICOM) consensus review on scientific advances & treatment paradigms for clinicians, researchers, and patients. Neuro Oncol. 2024.

7. Kshettry VR, Ostrom QT, Kruchko C, Al-Mefty O, Barnett GH, Barnholtz-Sloan JS. Descriptive epidemiology of World Health Organization grades II and III intracranial meningiomas in the United States. Neuro Oncol. 2015;17(8):1166–1173.

8. Louis DN, Perry A, Wesseling P, et al. The 2021 WHO Classification of Tumors of the Central Nervous System: a summary. Neuro Oncol. 2021;23(8):1231–1251.

9. Nassiri F, Liu J, Patil V, et al. A clinically applicable integrative molecular classification of meningiomas. Nature. 2021;597(7874):119–125.

10. Choudhury A, Magill ST, Eaton CD, et al. Meningioma DNA methylation groups identify biological drivers and therapeutic vulnerabilities. Nat Genet. 2022;54(5):649–659.

11. Sekely A, Zakzanis KK, Mabbott D, et al. Long-term neurocognitive, psychological, and return to work outcomes in meningioma patients. Support Care Cancer. 2022;30(5):3893–3902.

12. Central Brain Tumor Registry of the United States SEER*Stat Database. CDC National Program of Cancer Registries and NCI Surveillance, Epidemiology and End Results Incidence Data, 2020 submission (2000–2018). 2021.

13. SEER*Stat software [computer program]. Version version 8.4.02022.

14. Tiwari RC, Clegg LX, Zou Z. Efficient interval estimation for age-adjusted cancer rates. Stat Methods Med Res. 2006;15(6):547–569.

15. Walsh KM, Zhang C, Calvocoressi L, et al. Pleiotropic MLLT10 variation confers risk of meningioma and estrogen-mediated cancers. Neuro-Oncology Advances. 2022.

16. Magill ST, Vasudevan HN, Seo K, et al. Multiplatform genomic profiling and magnetic resonance imaging identify mechanisms underlying intratumor heterogeneity in meningioma. Nat Commun. 2020;11(1):4803.

17. Korhonen K, Salminen T, Raitanen J, Auvinen A, Isola J, Haapasalo H. Female predominance in meningiomas can not be explained by differences in progesterone, estrogen, or androgen receptor expression. J Neurooncol. 2006;80(1):1–7.

18. Pravdenkova S, Al-Mefty O, Sawyer J, Husain M. Progesterone and estrogen receptors: opposing prognostic indicators in meningiomas. J Neurosurg. 2006;105(2):163–173.

19. Hage M, Plesa O, Lemaire I, Raffin Sanson ML. Estrogen and Progesterone Therapy and Meningiomas. Endocrinology. 2022;163(2).

20. Bleil ME, Booth-LaForce C, Benner AD. Race disparities in pubertal timing: Implications for cardiovascular disease risk among African American women. Popul Res Policy Rev. 2017;36(5):717–738.

21. Takahashi H, Cornish AJ, Sud A, et al. Mendelian randomization provides support for obesity as a risk factor for meningioma. Sci Rep. 2019;9(1):309.

22. Schildkraut JM, Calvocoressi L, Wang F, et al. Endogenous and exogenous hormone exposure and the risk of meningioma in men. J Neurosurg. 2014;120(4):820–826.

23. Khazanchi R, Nandoliya KR, Shahin MN, et al. Obesity and meningioma: a US population-based study paired with analysis of a multi-institutional cohort. J Neurosurg. 2024;140(6):1558–1567.

24. Claus EB, Walsh KM, Calvocoressi L, et al. Cigarette smoking and risk of meningioma: the effect of gender. Cancer Epidemiol Biomarkers Prev. 2012;21(6):943–950.

25. Palmer JD, Prasad RN, Cioffi G, et al. Exposure to Radon and Heavy Particulate Pollution and Incidence of Brain Tumors. Neuro Oncol. 2022.

26. Bromberger JT, Matthews KA, Kuller LH, Wing RR, Meilahn EN, Plantinga P. Prospective study of the determinants of age at menopause. Am J Epidemiol. 1997;145(2):124–133.

27. McKnight KK, Wellons MF, Sites CK, et al. Racial and regional differences in age at menopause in the United States: findings from the REasons for Geographic And Racial Differences in Stroke (REGARDS) study. Am J Obstet Gynecol. 2011;205(4):353 e351–358.

28. Huttner HB, Bergmann O, Salehpour M, et al. Meningioma growth dynamics assessed by radiocarbon retrospective birth dating. EBioMedicine. 2018;27:176–181.

29. Chen WC, Choudhury A, Youngblood MW, et al. Targeted gene expression profiling predicts meningioma outcomes and radiotherapy responses. Nat Med. 2023;29(12):3067–3076.

30. Peng S, Cheng Z, Guo Z. Diagnostic nomogram model for predicting preoperative pathological grade of meningioma. Transl Cancer Res. 2021;10(9):4057–4064.

31. Han T, Liu X, Xu Z, et al. Preoperative Prediction of Meningioma Subtype by Constructing a Clinical-Radiomics Model Nomogram Based on Magnetic Resonance Imaging. World Neurosurg. 2024;181:e203–e213.

32. Herrgott GA, Snyder JM, She R, et al. Detection of diagnostic and prognostic methylation-based signatures in liquid biopsy specimens from patients with meningiomas. Nat Commun. 2023;14(1):5669.

