## Supplementary Tables and Figures for "Elevated meningioma risk among individuals who are Non-Hispanic Black is strongest for grade 2-3 tumors and synergistically modified by male sex"

**Supplementary Table 1.** Total cases, average annual age-adjusted incidence rate (AAAIR)[1], incidence rate ratio (IRR), and 95% confidence interval (CI) for meningioma by 10-year age intervals and stratified by biological sex (CBTRUS: Data provided by CDC's National Program of Cancer Registries and NCI's Surveillance, Epidemiology and End Results Program, 2004-2019).

| Sex | Age at Diagnosis | Race/Ethnicity | Total (2004-2019) | AAAIR (95% CI) | IRR (95% CI) | P-value |
|---|---|---|---|---|---|---|
| Male | 0-9 | White non-Hispanic | 118 | 0.07 (0.06-0.08) | REF. | |
| | 0-9 | Black non-Hispanic | 30 | 0.06 (0.04-0.09) | 0.93 (0.60-1.40) | 0.7922 |
| | 10-19 | White non-Hispanic | 422 | 0.21 (0.19-0.23) | REF. | |
| | 10-19 | Black non-Hispanic | 101 | 0.19 (0.15-0.23) | 0.90 (0.72-1.12) | 0.3657 |
| | 20-29 | White non-Hispanic | 1,311 | 0.65 (0.61-0.68) | REF. | |
| | 20-29 | Black non-Hispanic | 341 | 0.70 (0.63-0.78) | 1.09 (0.96-1.23) | 0.1801 |
| | 30-39 | White non-Hispanic | 3,335 | 1.71 (1.65-1.77) | REF. | |
| | 30-39 | Black non-Hispanic | 771 | 1.92 (1.78-2.06) | 1.12 (1.03-1.21) | **0.0053** |
| | 40-49 | White non-Hispanic | 7,370 | 3.26 (3.18-3.33) | REF. | |
| | 40-49 | Black non-Hispanic | 1,739 | 4.22 (4.02-4.42) | 1.30 (1.23-1.37) | **<0.0001** |
| | 50-59 | White non-Hispanic | 14,385 | 6.00 (5.90-6.10) | REF. | |
| | 50-59 | Black non-Hispanic | 3198 | 8.72 (8.41-9,02) | 1.45 (1.40-1.51) | **<0.0001** |
| | 60-69 | White non-Hispanic | 21,747 | 12.18 (12.02-12.35) | REF. | |
| | 60-69 | Black non-Hispanic | 3,583 | 16.50 (15.96-17.05) | 1.32 (1.27-1.37) | **<0.0001** |
| | 70-79 | White non-Hispanic | 22,948 | 21.199(21.71-22.28) | REF. | |
| | 70-79 | Black non-Hispanic | 2,983 | 29.06 (28.01-30.13) | 1.32 (1.27-1.37) | **<0.0001** |
| | 80+ | White non-Hispanic | 19,297 | 34.17 (33.69-34.66) | REF. | |
| | 80+ | Black non-Hispanic | 1,706 | 38.74 (36.91-40.62) | 1.13 (1.08-1.19) | **<0.0001** |
| Female | 0-9 | White non-Hispanic | 129 | 0.08 (0.06-0.09) | REF. | |
| | 0-9 | Black non-Hispanic | 18 | 0.04 (0.02-0.06) | 0.50 (0.29-0.82) | **0.0039** |
| | 10-19 | White non-Hispanic | 473 | 0.25 (0.23-0.27) | REF. | |
| | 10-19 | Black non-Hispanic | 119 | 0.23 (0.19-0.27) | 0.92 (0.75-1.13) | 0.4423 |
| | 20-29 | White non-Hispanic | 2,330 | 1.18 (1.13-1.23) | REF. | |
| | 20-29 | Black non-Hispanic | 731 | 1.47 (1.37-1.58) | 1.25 (1.14-1.35) | **<0.0001** |
| | 30-39 | White non-Hispanic | 9,910 | 5.19 (5.09-5.29) | REF. | |
| | 30-39 | Black non-Hispanic | 2,516 | 5.66 (5.44-5.89) | 1.09 (1.04-1.14) | **0.0001** |

| | | | | | | |
|---|---|---|---|---|---|---|
| | 40-49 | White non-Hispanic | 25,740 | 11.36 (11.22-11.50) | REF. | |
| | 40-49 | Black non-Hispanic | 5,840 | 12.58 (12.26-12.91) | 1.11 (1.08-1.14) | **<0.0001** |
| | 50-59 | White non-Hispanic | 44,123 | 18.07 (17.90-18.24) | REF. | |
| | 50-59 | Black non-Hispanic | 9,032 | 21.30 (20.867-21.75) | 1.18 (1.15-1.21) | **<0.0001** |
| | 60-69 | White non-Hispanic | 54,620 | 28.31 (28.07-28.55) | REF. | |
| | 60-69 | Black non-Hispanic | 9,490 | 34.42 (33.73-35.12) | 1.22 (1.19-1.24) | **<0.0001** |
| | 70-79 | White non-Hispanic | 53,728 | 42.84 (42.48-43.20) | REF. | |
| | 70-79 | Black non-Hispanic | 7,828 | 51.29 (50.16-52.44) | 1.20 (1.17-1.23) | **<0.0001** |
| | 80+ | White non-Hispanic | 52,554 | 54.73 (54.26-55.21) | REF. | |
| | 80+ | Black non-Hispanic | 5,986 | 64.06 (62.44-65.71) | 1.17 (1.14-1.20) | **<0.0001** |

[1]Rates are per 100,000 population and are age-adjusted to the 2000 US standard population.
Abbreviations: CBTRUS, Central Brain Tumor Registry of the United States.

**Supplementary Table 2.** Estimated contrasts from age-stratified interaction model.

| Age group | Level1 | Level2 | Ratio (95% CI) | p |
|---|---|---|---|---|
| 0-9 | (Female Non-Hispanic Black) | (Male Non-Hispanic Black) | 0.68 (0.32-1.42) | 0.664 |
| | (Female Non-Hispanic White) | (Female Non-Hispanic Black) | 1.86 (1.00-3.45) | 0.049 |
| | (Female Non-Hispanic White) | (Male Non-Hispanic Black) | 1.26 (0.75-2.11) | 0.664 |
| | (Female Non-Hispanic White) | (Male Non-Hispanic White) | 1.18 (0.85-1.64) | 0.664 |
| | (Male Non-Hispanic White) | (Female Non-Hispanic Black) | 1.58 (0.84-2.94) | 0.272 |
| | (Male Non-Hispanic White) | (Male Non-Hispanic Black) | 1.07 (0.63-1.80) | 0.742 |
| 10-19 | (Female Non-Hispanic Black) | (Male Non-Hispanic Black) | 1.19 (0.85-1.67) | 0.654 |
| | (Female Non-Hispanic White) | (Female Non-Hispanic Black) | 1.09 (0.85-1.41) | 0.882 |
| | (Female Non-Hispanic White) | (Male Non-Hispanic Black) | 1.31 (0.99-1.72) | 0.058 |
| | (Female Non-Hispanic White) | (Male Non-Hispanic White) | 1.17 (0.99-1.38) | 0.062 |
| | (Male Non-Hispanic White) | (Female Non-Hispanic Black) | 0.93 (0.72-1.21) | 0.882 |
| | (Male Non-Hispanic White) | (Male Non-Hispanic Black) | 1.12 (0.85-1.47) | 0.882 |
| 20-29 | (Female Non-Hispanic Black) | (Male Non-Hispanic Black) | 2.11 (1.78-2.50) | <0.001 |
| | (Female Non-Hispanic White) | (Female Non-Hispanic Black) | 0.82 (0.73-0.91) | <0.001 |
| | (Female Non-Hispanic White) | (Male Non-Hispanic Black) | 1.72 (1.48-2.00) | <0.001 |
| | (Female Non-Hispanic White) | (Male Non-Hispanic White) | 1.86 (1.71-2.04) | <0.001 |
| | (Male Non-Hispanic White) | (Female Non-Hispanic Black) | 0.44 (0.39-0.49) | <0.001 |
| | (Male Non-Hispanic White) | (Male Non-Hispanic Black) | 0.92 (0.79-1.08) | 0.185 |
| 30-39 | (Female Non-Hispanic Black) | (Male Non-Hispanic Black) | 2.93 (2.63-3.26) | <0.001 |
| | (Female Non-Hispanic White) | (Female Non-Hispanic Black) | 0.92 (0.87-0.98) | <0.001 |
| | (Female Non-Hispanic White) | (Male Non-Hispanic Black) | 2.70 (2.45-2.98) | <0.001 |
| | (Female Non-Hispanic White) | (Male Non-Hispanic White) | 3.00 (2.85-3.16) | <0.001 |
| | (Male Non-Hispanic White) | (Female Non-Hispanic Black) | 0.31 (0.29-0.33) | <0.001 |
| | (Male Non-Hispanic White) | (Male Non-Hispanic Black) | 0.90 (0.81-1.00) | 0.007 |
| 40-49 | (Female Non-Hispanic Black) | (Male Non-Hispanic Black) | 2.99 (2.78-3.21) | <0.001 |
| | (Female Non-Hispanic White) | (Female Non-Hispanic Black) | 0.91 (0.88-0.95) | <0.001 |
| | (Female Non-Hispanic White) | (Male Non-Hispanic Black) | 2.72 (2.55-2.90) | <0.001 |
| | (Female Non-Hispanic White) | (Male Non-Hispanic White) | 3.49 (3.37-3.61) | <0.001 |
| | (Male Non-Hispanic White) | (Female Non-Hispanic Black) | 0.26 (0.25-0.27) | <0.001 |
| | (Male Non-Hispanic White) | (Male Non-Hispanic Black) | 0.78 (0.73-0.83) | <0.001 |
| 50-59 | (Female Non-Hispanic Black) | (Male Non-Hispanic Black) | 2.47 (2.34-2.60) | <0.001 |
| | (Female Non-Hispanic White) | (Female Non-Hispanic Black) | 0.86 (0.83-0.89) | <0.001 |
| | (Female Non-Hispanic White) | (Male Non-Hispanic Black) | 2.12 (2.02-2.22) | <0.001 |
| | (Female Non-Hispanic White) | (Male Non-Hispanic White) | 3.01 (2.94-3.09) | <0.001 |
| | (Male Non-Hispanic White) | (Female Non-Hispanic Black) | 0.29 (0.28-0.30) | <0.001 |
| | (Male Non-Hispanic White) | (Male Non-Hispanic Black) | 0.70 (0.67-0.74) | <0.001 |
| 60-69 | (Female Non-Hispanic Black) | (Male Non-Hispanic Black) | 2.12 (2.02-2.23) | <0.001 |
| | (Female Non-Hispanic White) | (Female Non-Hispanic Black) | 0.84 (0.82-0.86) | <0.001 |
| | (Female Non-Hispanic White) | (Male Non-Hispanic Black) | 1.78 (1.70-1.86) | <0.001 |
| | (Female Non-Hispanic White) | (Male Non-Hispanic White) | 2.38 (2.34-2.43) | <0.001 |
| | (Male Non-Hispanic White) | (Female Non-Hispanic Black) | 0.35 (0.34-0.36) | <0.001 |
| | (Male Non-Hispanic White) | (Male Non-Hispanic Black) | 0.75 (0.71-0.78) | <0.001 |
| 70-79 | (Female Non-Hispanic Black) | (Male Non-Hispanic Black) | 1.80 (1.70-1.91) | <0.001 |
| | (Female Non-Hispanic White) | (Female Non-Hispanic Black) | 0.86 (0.83-0.89) | <0.001 |
| | (Female Non-Hispanic White) | (Male Non-Hispanic Black) | 1.55 (1.47-1.62) | <0.001 |
| | (Female Non-Hispanic White) | (Male Non-Hispanic White) | 2.00 (1.96-2.04) | <0.001 |
| | (Male Non-Hispanic White) | (Female Non-Hispanic Black) | 0.43 (0.41-0.44) | <0.001 |
| | (Male Non-Hispanic White) | (Male Non-Hispanic Black) | 0.77 (0.74-0.81) | <0.001 |
| 80+ | (Female Non-Hispanic Black) | (Male Non-Hispanic Black) | 1.66 (1.54-1.78) | <0.001 |
| | (Female Non-Hispanic White) | (Female Non-Hispanic Black) | 0.87 (0.84-0.90) | <0.001 |
| | (Female Non-Hispanic White) | (Male Non-Hispanic Black) | 1.44 (1.35-1.54) | <0.001 |
| | (Female Non-Hispanic White) | (Male Non-Hispanic White) | 1.62 (1.59-1.66) | <0.001 |
| | (Male Non-Hispanic White) | (Female Non-Hispanic Black) | 0.54 (0.52-0.56) | <0.001 |
| | (Male Non-Hispanic White) | (Male Non-Hispanic Black) | 0.89 (0.83-0.95) | <0.001 |

**Supplementary Table 3.** Total cases, average annual age-adjusted incidence rate (AAAIR), incidence rate ratio (IRR), and 95% confidence interval (CI) for meningioma by 10-year age intervals and stratified by central nervous system (CNS) WHO grade (CBTRUS: Data provided by CDC's National Program of Cancer Registries and NCI's Surveillance, Epidemiology and End Results Program, 2004-2019).

| CNS WHO grade | Age at Diagnosis | Race/Ethnicity | Total (2004-2019) | AAAIR (95% CI) | IRR (95% CI) | P-value |
|---|---|---|---|---|---|---|
| Grade 1 | 0-9 | White non-Hispanic | 160 | 0.05 (0.04-0.05) | REF. | |
| | | Black non-Hispanic | 35 | 0.04 (0.03-0.05) | 0.79 (0.53-1.14) | 02288 |
| | 10-19 | White non-Hispanic | 705 | 0.18 (0.17-0.19) | REF. | |
| | | Black non-Hispanic | 168 | 0.16 (0.14-0.18) | 0.88 (0.74-1.04) | 0.1493 |
| | 20-29 | White non-Hispanic | 3,148 | 0.79 (0.76-0.82) | REF. | |
| | | Black non-Hispanic | 958 | 0.98 (0.92-1.05) | 1.25 (1.16-1.33) | **<0.0001** |
| | 30-39 | White non-Hispanic | 12,090 | 3.14 (3.08-3.20) | REF. | |
| | | Black non-Hispanic | 3,000 | 3.55 (3.42-3.68) | 1.13 (1.09-1.18) | **<0.0001** |
| | 40-49 | White non-Hispanic | 30,902 | 6.82 (6.74-6.90) | REF. | |
| | | Black non-Hispanic | 6,950 | 7.92 (7.74-8.11) | 1.16 (1.13-1.19) | **<0.0001** |
| | 50-59 | White non-Hispanic | 54,868 | 11.36 (11.27-11.46) | REF. | |
| | | Black non-Hispanic | 11,377 | 14.39 (14.13-14.66) | 1.27 (1.24-1.29) | **<0.0001** |
| | 60-69 | White non-Hispanic | 71,755 | 19.31 (19.17-19.45) | REF. | |
| | | Black non-Hispanic | 12,115 | 24.58 (24.14-25.03) | 1.27 (1.25-1.30) | **<0.0001** |
| | 70-79 | White non-Hispanic | 72,673 | 31.61 (31.38-31.84) | REF. | |
| | | Black non-Hispanic | 10,136 | 39.72 (38.94-40.50) | 1.26 (1.23-1.28) | **<0.0001** |
| | 80+ | White non-Hispanic | 69,484 | 45.38 (45.04-45.72) | REF. | |
| | | Black non-Hispanic | 7,412 | 53.89 (52.67-55.13) | 1.19 (1.16-1.22) | **<0.0001** |
| Grades 2-3 | 0-9 | White non-Hispanic | 87 | 0.03 (0.02-0.03) | REF. | |
| | | Black non-Hispanic | <16 cases | -- | -- | -- |
| | 10-19 | White non-Hispanic | 190 | 0.05 (0.04-0.06) | REF. | |

| | | | | | | |
|---|---|---|---|---|---|---|
| | | Black non-Hispanic | 52 | 0.05 (0.04-0.07) | 1.02 (0.74-1.39) | 0.9522 |
| | 20-29 | White non-Hispanic | 493 | 0.12 (0.11-0.13) | REF. | |
| | | Black non-Hispanic | 114 | 0.11 (0.09-0.14) | 0.93 (0.75-1.14) | 0.4943 |
| | 30-39 | White non-Hispanic | 1,155 | 0.30 (0.28-0.32) | REF. | |
| | | Black non-Hispanic | 287 | 0.34 (0.30-0.38) | 1.14 (1.00-1.30) | 0.0590 |
| | 40-49 | White non-Hispanic | 2,208 | 0.49 (0.47-0.51) | REF. | |
| | | Black non-Hispanic | 629 | 0.73 (0.67-0.78) | 1.48 (1.35-1.62) | **<0.0001** |
| | 50-59 | White non-Hispanic | 3,640 | 0.75 (0.73-0.78) | REF. | |
| | | Black non-Hispanic | 853 | 1.07 (1.00-1.15) | 1.42 (1.32-1.54) | **<0.0001** |
| | 60-69 | White non-Hispanic | 4,612 | 1.24 (1.20-1.28) | REF. | |
| | | Black non-Hispanic | 958 | 1.94 (1.82-2.07) | 1.57 (1.46-1.69) | **<0.0001** |
| | 70-79 | White non-Hispanic | 4,003 | 1.74 (1.68-1.79) | REF. | |
| | | Black non-Hispanic | 675 | 2.62 (2.43-2.83) | 1.51 (1.39-1.64) | **<0.0001** |
| | 80+ | White non-Hispanic | 2,367 | 1.57 (1.51-1.64) | REF. | |
| | | Black non-Hispanic | 280 | 2.04 (1.81-2.30) | 1.30 (1.14-1.47) | **0.0001** |

**Supplementary Figure 1.** Non-Hispanic Black to non-Hispanic White incidence rate ratios (IRRs) and 95% confidence intervals (CI) for meningioma, stratified by 10-year age group at diagnosis and surgery status. (CBTRUS: Data provided by CDC's National Program of Cancer Registries and NCI's Surveillance, Epidemiology and End Results Program, 2004-2019).

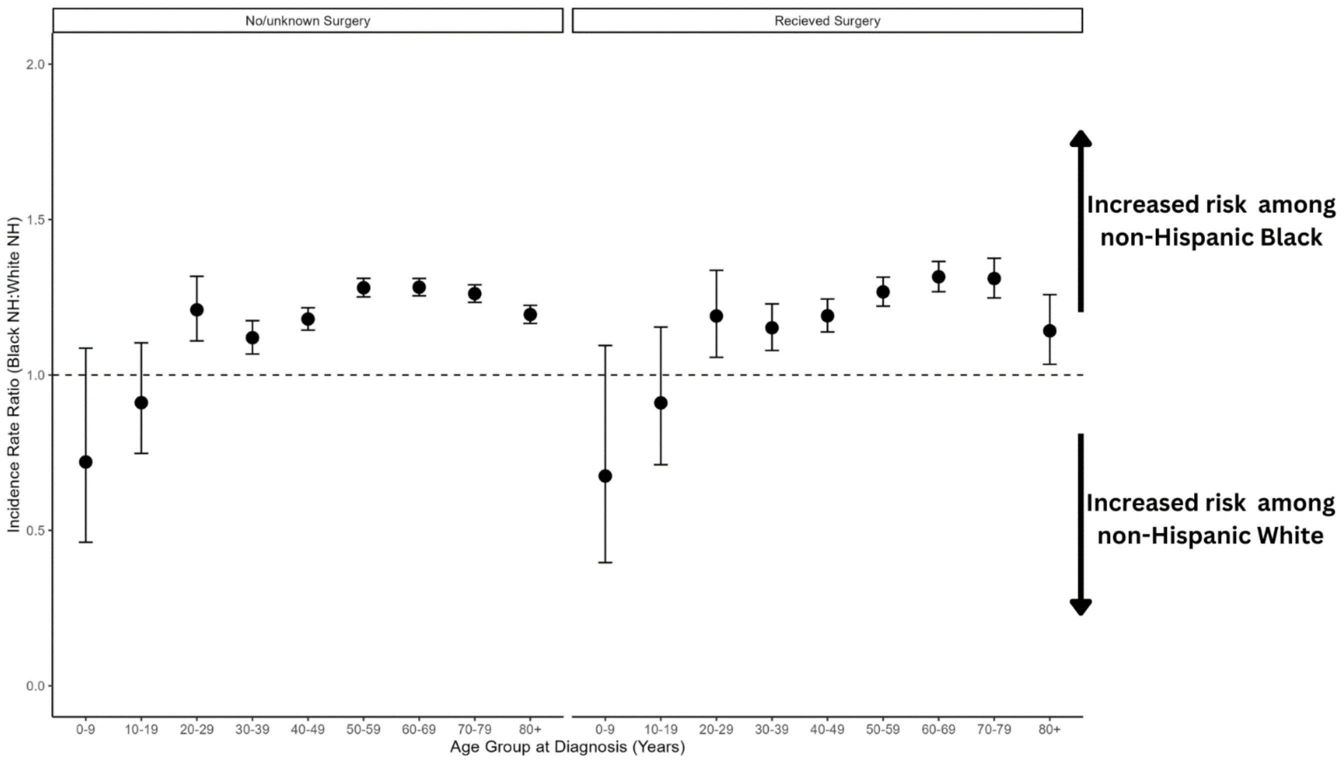

**Supplementary Figure 2.** Non-Hispanic Black to non-Hispanic White incidence rate ratios (IRRs) and 95% confidence intervals (CI) for meningioma, by 10-year age group at diagnosis and stratified by sex and by central nervous system (CNS) WHO grade (CBTRUS: Data provided by CDC's National Program of Cancer Registries and NCI's Surveillance, Epidemiology and End Results Program, 2004-2019). Asterisks denote statistically significant interaction (P<0.05) in the meningioma risk associated with race/ethnicity among males compared to females (CBTRUS: Data provided by CDC's National Program of Cancer Registries and NCI's Surveillance, Epidemiology and End Results Program, 2004-2019).

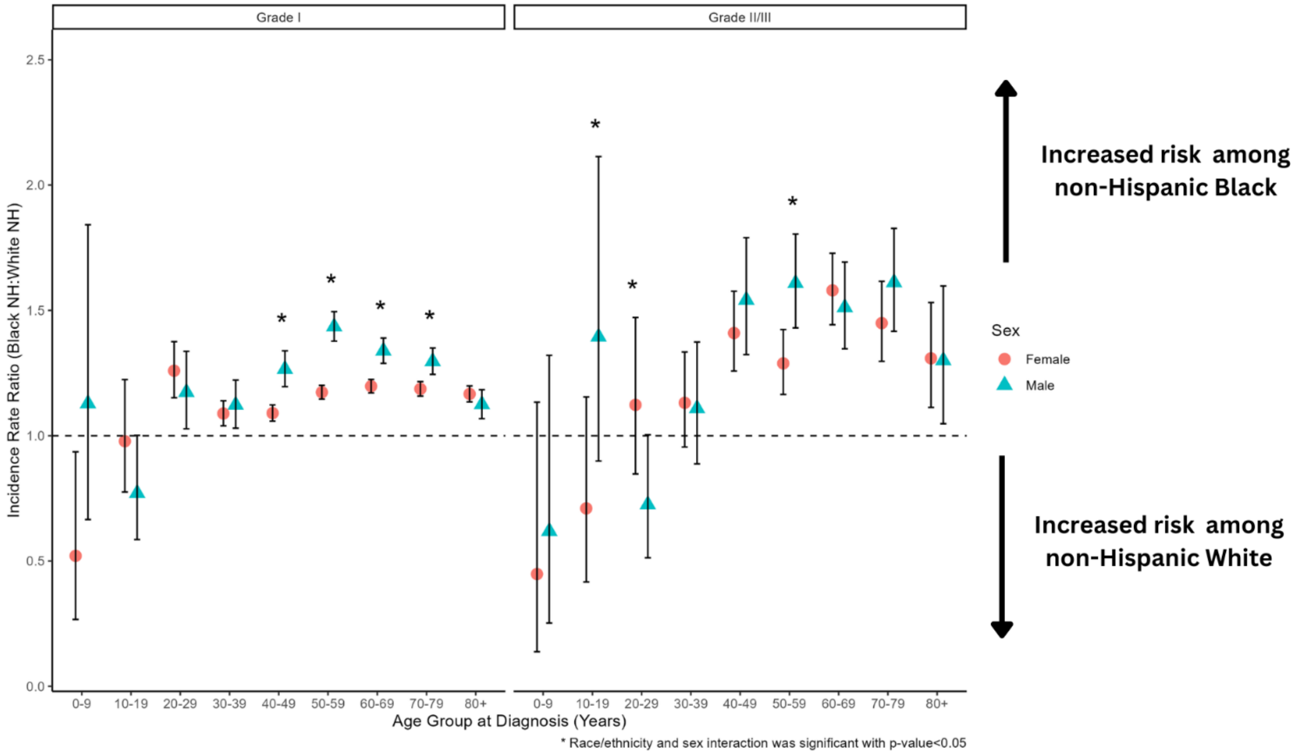
